# Risk factors and cognitive domain markers of progression in subjective cognitive decline

**DOI:** 10.1101/2024.08.22.24312451

**Authors:** Omonigho M. Bubu, Alfred K. Mbah, Mark A. Bernard, Anthony Q. Briggs, Arline Faustin, Lindsey Gurin, Julia A. Rao, Sakina Ouedraogo Tall, Ricardo S. Osorio, Arjun V. Masurkar

## Abstract

We evaluated risk factor differences and cognitive domain markers associated with progression in subjects with subjective cognitive decline (SCD) at baseline from the NYU Alzheimer’s Disease Research Center. We included SCD non-decliners (n = 27), who remained stable, and decliners (n = 24), who progressed to mild cognitive impairment or worse, between the second to sixth yearly follow-up visits. Adjusted mixed-effects models examined group differences and associations between demographic, APOE status, psychometric test performance and comorbidities with longitudinal-decline. Overall, mean (SD) age was 67.4 (9.2) and total follow-up time was 5.1 (1.8) years. Lower education (14.9 (3.2) vs. 17.3 (2.1)), Hispanic ethnicity (50.0% vs. 11.0%), and hypercholesterolemia (aOR: 6.67), *p* ≤ 0.05 for all, were risk factors for progression in SCD, whereas APOE status was not. Notably, SCD decliners were at increased risk for both amnestic and non-amnestic cognitive-decline with psychometric changes in memory, executive, and language domains (*p* < 0.001 for all). These findings inform further work on SCD outcomes and related biomarkers, as well as preventive studies that target modifiable risk factors for SCD progression.

## Introduction

Subjective cognitive decline (SCD) is considered a preclinical stage of Alzheimer’s disease (AD), identifying persons with normal performance on psychometric tests who progress to subsequent AD stages faster than cognitively normal individuals without subjective complaints.^1^ Consequently, SCD also associates with imaging and biofluids biomarkers of AD^2-5^.

Specifically, previous work from our center^6^ showed that SCD is a precursor of further decline to mild cognitive impairment (MCI) or dementia over a 7-year mean follow-up interval. Specifically, in a sample of 260 subjects (166 cognitively normal with SCD and 47 cognitively normal without SCD), SCD subjects were 4.5 times more likely to decline to mild cognitive impairment (MCI) or dementia. We subsequently studied features of decline in 98 SCD subjects over a 2-year interval, noting worsening scores on the Digit Symbol Substitution Test and the Brief Cognitive Rating Scale for staging assessment, as well as a reduction in anxiety^7^.

Subsequently, we reported on the characteristics of a pre-SCD condition materializing in likely AD. In this study 47 cognitively normal healthy older subjects (mean baseline age= 64.1±8.9 years) were followed for approximately 6.7±3.1 years. After controlling for age, sex, years of education, and follow-up time, there were significant group differences in the rate of decline and composite psychometric scores between future decliners (n=36) relative to non-decliners (n=11).^8^

As such, SCD can be a precursor to MCI or AD dementia, taking as long as 7 to 15 years for transition to occur. However, stage transition rates may differ even among SCD subjects, as we have found in the pre-SCD interval^8^. Moreover, the specific drivers and nature of SCD progression remain unclear. Therefore, in this study we evaluated risk factor differences between SCD decliners and non-decliners related to demographics, genetics, and presence of comorbidities. We also investigated cognitive domain markers associated with progression over time in order to investigate the nature of progression in subjects with SCD.

## Methods

### Study Design, Participants and Variables

#### Study Design

A retrospective longitudinal analysis was conducted on selected subjects followed at the NYU Alzheimer’s Disease Research Center (ADRC). The NYU Grossman School of Medicine Institutional Review Board approved all aspects of this study, and written informed consent was obtained from all subjects.

#### Participants

Subjects were community dwelling, over 55 years of age, recruited by public announcement or referral to participate in this longitudinal study on cognition and brain aging. Inclusion for the ADRC include (1) having a study partner, (2) fluency in English and Spanish (including their study partner), and (3) willingness to undergo ADRC Clinical Core evaluation that includes imaging and blood draw. Exclusion criteria include (1) history of moderate/severe psychiatric illness, significant neurological disease other than AD and related disorders, significant organ disease or transplant, autoimmune disease requiring treatment; malignancy within 5 years (other than low grade prostate cancer, non-melanotic skin cancer); (2) alcohol or drug abuse. Participants are followed yearly, and the ADRC Clinical Core evaluations include full clinical, neurological, psychiatric, neuropsychological, clinical laboratory, and neuroimaging evaluations according to the National Alzheimer’s Coordinating Center Uniform Data Set 3.0. For our analysis we analyzed a subset of Clinical Core participants that (1) had a consensus diagnosis of SCD at baseline by the Global Deterioration Scale^9^ and (2a) who remained cognitively and functionally normal for six annual visits, deemed “non-decliners” or (2b) who progressed to MCI or dementia between the second to sixth follow-up visits, deemed “decliners.” This resulted in an analysis cohort of 27 non-decliners and 24 decliners. Subjects included in this analysis cohort were recruited between January 1, 2013 and December 31, 2015, followed at yearly intervals until December 31, 2021. Follow-up evaluations were completed without reference to the prior results.

#### Demographic variables

Demographic variables included age (calculated from reported date of birth), sex (biological variable as reported by subject), years of education (calculated based on highest reported completed diploma/degree), and body mass index (calculated from measured weight and height).

#### Genetic variable (APOE)

APOE genotype was characterized as having at least one APOE □4 allele or at least one APOE □2 allele.

#### Medical Co-morbidities

Clinical co-morbidities assessed included clinical history of hypertension, diabetes, anxiety, insomnia, obstructive sleep apnea, osteoarthritis, hypercholesterolemia, and thyroid disease. These conditions were reported by subjects during clinical assessment.

#### Cognitive variables

Cognitive measures assessed in this study included the functional and cognitive assessments used in the National Alzheimer’s Coordinating Center’s Uniform Data Set 3.0: Clinical Dementia Rating (CDR) scale, Mini Mental State Exam (MMSE), Montreal Cognitive Assessment (MoCA), Digit Span Recall (Forwards, Backwards), Benton Visual Retention Tests, Craft story, Multilingual Naming Test (MINT), Vegetable and Animal naming, Trail Making Test Part A and Part B.

### Statistical Analysis

All models included age, sex, education, self-reported Hispanic ethnicity, baseline psychometric data, and APOE genotype.

#### Comparison of baseline participant characteristics between SCD decliners vs. non decliners

Generalized linear models examined group differences in continuous demographic risk factors. Differences in categorical variables were assessed with the Fisher exact test to compare proportions of subjects between SCD groups.

#### Comparison of Psychometric test performance over time in SCD decliners vs. non-decliners

Analysis of Covariance (ANCOVA) was conducted to test whether significant differences in changes in mean cognitive scores existed between SCD groups over time. In the ANCOVA analysis, time was treated as continuous. Psychometric test performance associations with longitudinal decline, comparing decliners to non-decliners, were also investigated with linear mixed effects models.

#### Examining associations of medical comorbidities with longitudinal decline

To investigate the associations of medical comorbidities with longitudinal decline, comparing decliners to non-decliners, logistic (i.e., non-linear) mixed-effects models with random intercept and slope were used controlling for covariates/potential confounders, and their interactions with time (operationalized as years from baseline for each participant).

## Results

### Comparison of baseline participant characteristics between SCD decliners vs. non decliners

At baseline, there were no significant differences between decliners and non-decliners in age, sex, clinical co-morbidities, APO-ε2 or APO-ε4 carrier status (Table 1). The mean (SD) age for decliners vs. non-decliners was 68.5 (11.3) vs. 66.3 (7.1) years (p =0.41). The proportion of females was 67% vs. 74%, and APOE4 was 29% vs. 33%, *p*≥0.70 for both. There were significant differences in follow-up time, years of education and Hispanic ethnicity. The mean (SD follow-up time was 3.7 (1.97) vs. 6.3 (1.60) years, (*p* <0.001) for decliners vs. non-decliners. Subjects with lower years of education (14.9 (3.2) vs. 17.3 (2.1)) and those of Hispanic ethnicity (50% vs. 11%) were more likely to be decliners (*p* ≤ 0.004 for both).

**Table 1:** GLM Comparison of Participant characteristics at Baseline of Subjective Cognitive Decline (SCD) Groups: Decliners vs. Non-Decliners, NYU Alzheimer’s Disease Research Center.

| Characteristics | All<br>n=51 | SCD Group |  | P-value |
| --- | --- | --- | --- | --- |
|  |  | Decliners<br>(n=24) | Non-Decliners<br>(n=27) |  |
| Age, mean (SD), y | 67.4 (9.2) | 68.5 (11.3) | 66.3 (7.1) | 0.40 |
| Average Follow-up time, mean (SD), y | 5.1 (1.8) | 3.7 (2.0) | 6.3 (1.6) | <.001 |
| Female, n *(%) | 36 (71) | 16 (67) | 20 (74) | 0.72 |
| Male, n *(%) | 15 (29) | 8 (33) | 7(26) |  |
| Education, years (SD) | 16.1 (4.3) | 14.9 (3.2) | 17.3 (2.1) | 0.003 |
| Clinical co-morbid conditions |  |  |  |  |
| Hypertension n *(%) | 23 (45) | 13 (54) | 14 (52) | 0.42 |
| SBP mm, Hg mean (SD), | 141.5 (17.6) | 142.8 (18.0) | 139.8 (16.2) | 0.33 |
| Diabetes n *(%) | 6 (12) | 3 (13) | 3 (11) | 0.44 |
| Body mass index, mean ± SD | 27.7 ± 4.8 | 27.9 ± 5.8 | 27.4 ± 5.0 | 0.23 |
| Current smoker, n *(%) | 4 (8) | 2 (8) | 2 (7) | 0.76 |
| Anxiety, n *(%) | 5 (10) | 2 (8) | 3 (11) | 0.73 |
| Atrial Fibrillation, n *(%) | 6 (12) | 2(8) | 4 (15) | 0.67 |
| Hypercholesterolemia, n *(%) | 23 (45) | 18 (75) | 5 (19) | 0.42 |
| Osteoarthritis, n *(%) | 6 (12) | 2(8) | 4 (15) | 0.89 |
| Hyposomnia, n *(%) | 7 (14) | 3 (13) | 4 (15) | 0.43 |
| Sleep apnea, n *(%) | 4 (8) | 2 (8) | 2 (7) | 0.99 |
| Thyroid Disease, n *(%) | 16 (31) | 8 (30) | 8 (30) | 0.18 |
| Average number of vascular risk factors (SD) | 1.5 (1.1) | 1.5 (1.3) | 1.3 (1.1) | 0.23 |
| APOE genotype |  |  |  |  |
| Having at least one APOE ε4 allele, n *(%) | 15 (28) | 7 (29) | 9 (33) | 0.84 |
| Having at least one APOE ε2 allele, n *(%) | 6 (12) | 1 (4) | 5 (19) | 0.13 |
| Race and Ethnicity n *(%) |  |  |  |  |
| Black/African-American | 5 (10) | 4 (17) | 1 (4) | 0.14 |
| White | 44 (86) | 18 (75) | 26 (96) |  |
| Asian | 2 (4) | 2 (8) | 0 (0) |  |
| Hispanic Ethnicity n *(%) |  |  |  |  |
| No | 36 (71) | 12 (50) | 24 (89) | 0.004 |
| Yes | 15 (29) | 12 (50) | 3 (11) |  |
| MMSE median (interquartile range) | 29 (28, 30) | 29 (27, 30) | 29 (27, 30) | 0.99 |
| CDR median (interquartile range) | 0 (0, 0) | 0 (0, 0) | 0 (0, 0) | 0.99 |

\*(%), represents column percent; APOE represents apolipoprotein E, GLM represents generalized linear models; mean (SD) represents mean (standard deviation), MMSE represents mini mental state examination; n represents sample size, SBP represents systolic blood pressure; SD, standard deviation. Significant *P*-value $\leq 0.05$ .

### Comparison of Psychometric test performance over time in SCD decliners vs. non-decliners

Results of the ANCOVA analysis are shown in Table 2. SCD decliners were at increased risk for psychometric changes in the Craft story (verbal short-term memory), Multilingual Naming Test (MINT), Vegetable and Animal semantic fluency tests, and the Trail Making Test Part A and Part B assessing for processing speed and executive function (*p* < 0.001 for all).

**Table 2:**
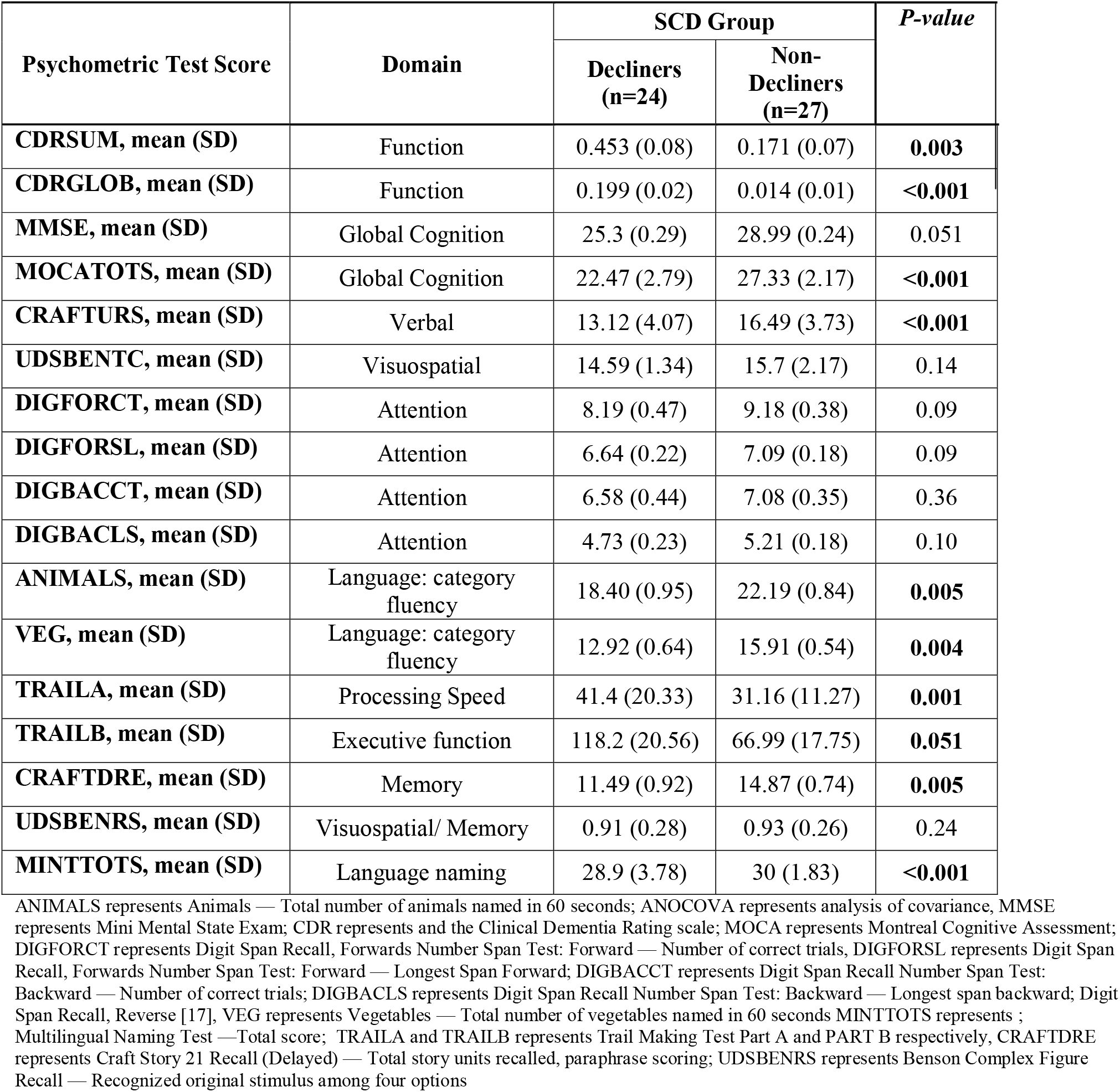
ANCOVA Comparisons of Psychometric test performance over time in SCD decliners vs. non-decliners.

### Examining associations of medical comorbidities with longitudinal decline

Logistic mixed effects estimates examining associations of medical comorbidities with longitudinal decline are shown in Figure 1. Only hypercholesteremia had a significant association. Decliners were approximately 7 times more likely (aOR: 6.67; 95% CI: 1.30 – 33.33, *p* =0.002) to have hypercholesterolemia compared to non-decliners.

**Figure 1:**
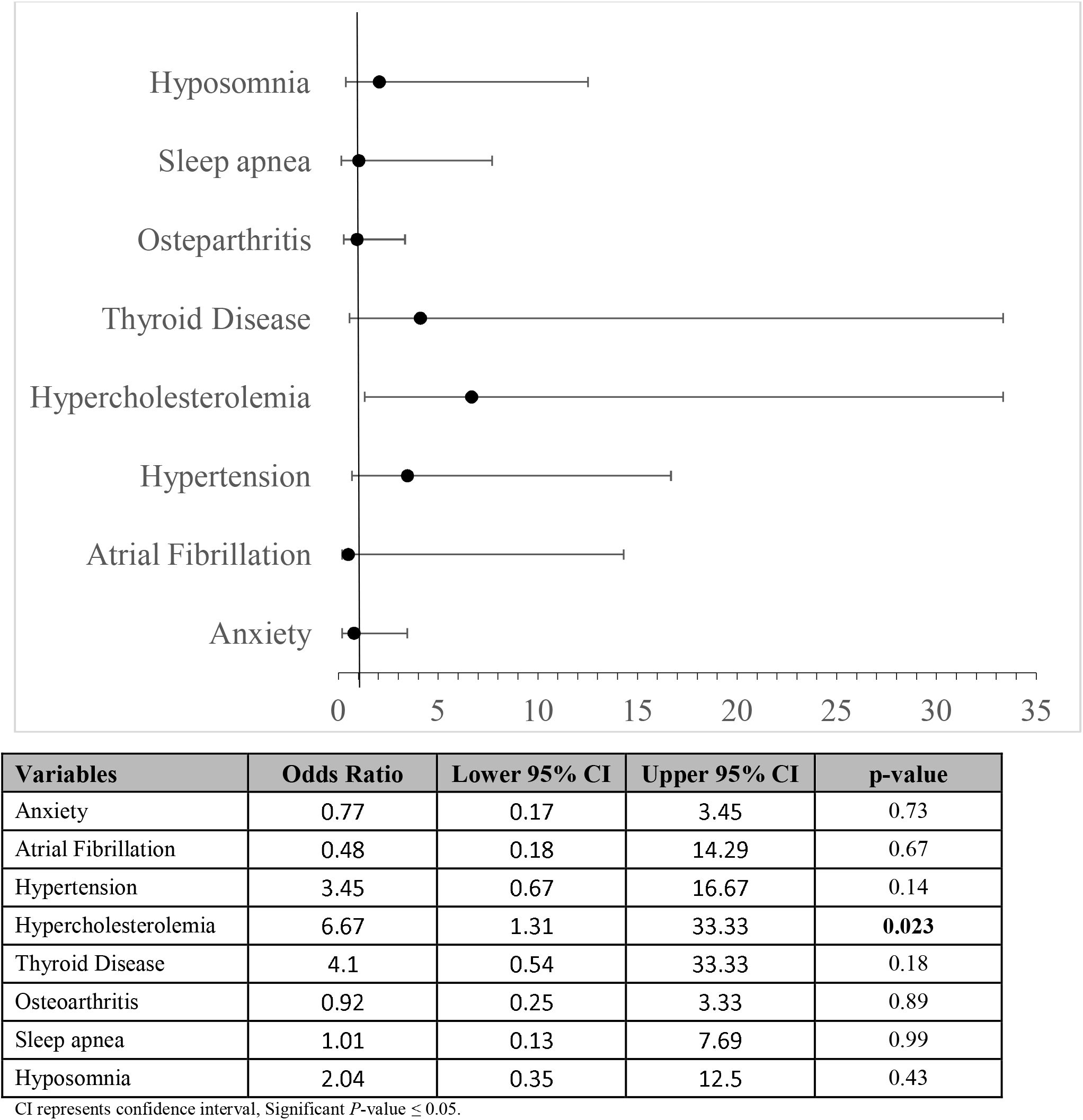
Logistic Mixed Effects Model Showing Effect Sizes Comparing Decliners vs. Non-Decliners over Time.

## Discussion

Our findings indicate that lower education, Hispanic ethnicity, and hypercholesterolemia are risk factors for progression in SCD, whereas APOE-ε4 carriage is not. The nature of progression showed prospective decline in psychometric tests across multiple domains: memory, language (naming and semantic fluency), processing speed and executive function. Thus, SCD decliners are at increased risk for both amnestic and non-amnestic cognitive decline.

Regarding the demographics risks factors, education and related factors of educational quality and literacy are well established risk factors for dementia.^10,11^ Racial and ethnic minorities (e.g. Hispanic persons) have disproportionately higher rates of dementia and clinical AD,^12^ and vascular risk factors (e.g. hypercholesterolemia) lower the threshold for cognitive impairment.^13,14^ Moreover, such factors are synergistic in that racial and ethnic minorities with lower education, and associated social determinants of low income and reduced health care, have even higher risk of SCD^15^. Additionally, these challenges can be exacerbated when individuals fail to discuss cognitive decline with health care professionals. Thus, our results suggest that SCD screening, monitoring, and intervention in these groups is of high clinical importance.

Our finding of hypercholesterolemia as a risk factor for SCD progression is not only expected as a risk factor for vascular cognitive impairment, but it is also has links to AD. Indeed, we have previously observed that hyperlipidemia is equivalently prevalent in MCI from neuropathologically-confirmed AD and microvascular disease^16,17^. Yet, imaging biomarkers of microvascular disease (e.g. white matter hyperintensities on MRI FLAIR sequence) are not driven by hypercholesterolemia in SCD^18^. However, hypercholesterolemia has been associated with elevated brain amyloid levels and hypometabolism in AD-susceptible brain regions^19-22^. As it is a modifiable factor, lipid control in persons with SCD may be an important intervention to prevent the onset of objective cognitive decline.

APOE-ε4 carriage was surprisingly not a risk factor for progression in SCD in our cohort, despite its well-known association with increase AD risk^23^ likely due to increased amyloid deposition^24^. In other studies of SCD, APOE-ε4 carriage has been associated with higher CSF p-tau, lower CSF AB42, and higher amyloid deposition^25^. Such abnormalities, particularly CSF AB42/tau ratio, are reliably predictive of SCD progression to MCI and AD dementia^26^. One possibility to explain this discrepancy, apart from cohort and SCD assessment differences, is that SCD progression may correlate with accelerated spread of tauopathy, which may be less driven by APOE-ε4.

Furthermore, since we do not have biomarkers assessments in this cohort, it is also possible that some of the progression is due to vascular disease. This may be supported by aspects of non-amnestic progression and hyperlipidemia as a risk factor. Future work could potentially observe biofluids and/or imaging markers of amyloid, tau, and cerebrovascular disease in SCD participants to investigate their predictive value for amnestic vs. non-amnestic decline. Such work would help establish these biomarkers as complements to psychometric testing as predictors of disease status and progression, and provide more objective clinical insights into SCD pathology.

Strengths of our study include the comprehensive psychometric assessments and the longitudinal follow-up of study participants. Limitations of this study include the small sample size, as well as the absence of established markers of AD neuropathology such as amyloid and tau.

In summary, our findings inform on the design of biomarker studies of SCD outcomes, as well as preventive studies of MCI and eventual AD that target education and vascular risk in SCD subjects.

## Data Availability

All data produced in the present study are available upon reasonable request to the authors

## Acknowledgements

This work was funded by NIH P30 AG008051 and P30 AG066512.

## References

1 Rabin, L. A., Smart, C. M. & Amariglio, R. E. Subjective Cognitive Decline in Preclinical Alzheimer’s Disease. Annual review of clinical psychology 13, 369–396, doi:10.1146/annurev-clinpsy-032816-045136 (2017).

2 Buckley, R. F. et al. Using subjective cognitive decline to identify high global amyloid in community-based samples: A cross-cohort study. Alzheimer’s & Dementia: Diagnosis, Assessment & Disease Monitoring 11, 670–678, doi:10.1016/j.dadm.2019.08.004 (2019).

3 Wolfsgruber, S. et al. Prevalence of abnormal Alzheimer’s disease biomarkers in patients with subjective cognitive decline: cross-sectional comparison of three European memory clinic samples. Alzheimer’s Research & Therapy 11, 8, doi:10.1186/s13195-018-0463-y (2019).

4 Amariglio, R. E. et al. Subjective cognitive complaints and amyloid burden in cognitively normal older individuals. Neuropsychologia 50, 2880–2886, doi:10.1016/j.neuropsychologia.2012.08.011 (2012).

5 Hu, X. et al. Smaller medial temporal lobe volumes in individuals with subjective cognitive decline and biomarker evidence of Alzheimer’s disease—Data from three memory clinic studies. Alzheimer’s & Dementia 15, 185–193, doi:10.1016/j.jalz.2018.09.002 (2019).

6 Reisberg, B., Shulman, M. B., Torossian, C., Leng, L. & Zhu, W. Outcome over seven years of healthy adults with and without subjective cognitive impairment. Alzheimer’s & dementia : the journal of the Alzheimer’s Association 6, 11–24, doi:10.1016/j.jalz.2009.10.002 (2010).

7 Reisberg, B. et al. Two Year Outcomes, Cognitive and Behavioral Markers of Decline in Healthy, Cognitively Normal Older Persons with Global Deterioration Scale Stage 2 (Subjective Cognitive Decline with Impairment). J Alzheimers Dis 67, 685–705, doi:10.3233/JAD-180341 (2019).

8 Reisberg, B. et al. Psychometric Cognitive Decline Precedes the Advent of Subjective Cognitive Decline in the Evolution of Alzheimer’s Disease. Dement Geriatr Cogn Disord 49, 16–21, doi:10.1159/000507286 (2020).

9 Reisberg, B., Ferris, S. H., de Leon, M. J. & Crook, T. The Global Deterioration Scale for assessment of primary degenerative dementia. Am J Psychiatry 139, 1136–1139, doi:10.1176/ajp.139.9.1136 (1982).

10 Lövdén, M., Fratiglioni, L., Glymour, M. M., Lindenberger, U. & Tucker-Drob, E. M. Education and Cognitive Functioning Across the Life Span. Psychological science in the public interest : a journal of the American Psychological Society 21, 6–41, doi:10.1177/1529100620920576 (2020).

11 Sisco, S. et al. The role of early-life educational quality and literacy in explaining racial disparities in cognition in late life. The journals of gerontology. Series B, Psychological sciences and social sciences 70, 557–567, doi:10.1093/geronb/gbt133 (2015).

12 Chen, C. & Zissimopoulos, J. M. Racial and ethnic differences in trends in dementia prevalence and risk factors in the United States. Alzheimer’s & dementia (New York, N. Y.) 4, 510–520, doi:10.1016/j.trci.2018.08.009 (2018).

13 Esiri, M. M., Nagy, Z., Smith, M. Z., Barnetson, L. & Smith, A. D. Cerebrovascular disease and threshold for dementia in the early stages of Alzheimer’s disease. Lancet (London, England) 354, 919–920, doi:10.1016/s0140-6736(99)02355-7 (1999).

14 Schneider, J. A., Boyle, P. A., Arvanitakis, Z., Bienias, J. L. & Bennett, D. A. Subcortical infarcts, Alzheimer’s disease pathology, and memory function in older persons. Annals of neurology 62, 59–66, doi:10.1002/ana.21142 (2007).

15 Gupta, S. Racial and ethnic disparities in subjective cognitive decline: a closer look, United States, 2015-2018. BMC Public Health 21, 1173, doi:10.1186/s12889-021-11068-1 (2021).

16 Yang, D. & Masurkar, A. Early-Stage MRI Volumetric Differences in White Matter Hyperintensity and Temporal Lobe Volumes between Autopsy-Confirmed Alzheimer’s Disease, Cerebral Small Vessel Disease, and Mixed Pathologies. Dement Geriatr Cogn Dis Extra 12, 69–75, doi:10.1159/000524499 (2022).

17 Yang, D. & Masurkar, A. V. Clinical Profiles of Arteriolosclerosis and Alzheimer Disease at Mild Cognitive Impairment and Mild Dementia in a National Neuropathology Cohort. Alzheimer Dis Assoc Disord 35, 14–22, doi:10.1097/WAD.0000000000000411 (2021).

18 Rothstein, A. et al. Impact of white matter hyperintensities on subjective cognitive decline phenotype in a diverse cohort of cognitively normal older adults. Int J Geriatr Psychiatry 38, e5948, doi:10.1002/gps.5948 (2023).

19 Kobe, T. et al. Association of Vascular Risk Factors With beta-Amyloid Peptide and Tau Burdens in Cognitively Unimpaired Individuals and Its Interaction With Vascular Medication Use. JAMA Netw Open 3, e1920780, doi:10.1001/jamanetworkopen.2019.20780 (2020).

20 Pappolla, M. A. et al. Mild hypercholesterolemia is an early risk factor for the development of Alzheimer amyloid pathology. Neurology 61, 199–205, doi:10.1212/01.wnl.0000070182.02537.84 (2003).

21 Reed, B. et al. Associations between serum cholesterol levels and cerebral amyloidosis. JAMA Neurol 71, 195–200, doi:10.1001/jamaneurol.2013.5390 (2014).

22 Reiman, E. M. et al. Higher serum total cholesterol levels in late middle age are associated with glucose hypometabolism in brain regions affected by Alzheimer’s disease and normal aging. Neuroimage 49, 169–176, doi:10.1016/j.neuroimage.2009.07.025 (2010).

23 Corder, E. H. et al. Gene dose of apolipoprotein E type 4 allele and the risk of Alzheimer’s disease in late onset families. Science (New York, N.Y.) 261, 921–923 (1993).

24 Reiman, E. M. et al. Fibrillar amyloid-beta burden in cognitively normal people at 3 levels of genetic risk for Alzheimer’s disease. Proc Natl Acad Sci U S A 106, 6820–6825, doi:10.1073/pnas.0900345106 (2009).

25 Risacher, S. L. et al. APOE effect on Alzheimer’s disease biomarkers in older adults with significant memory concern. Alzheimer’s & Dementia 11, 1417–1429, doi:10.1016/j.jalz.2015.03.003 (2015).

26 Wolfsgruber, S. et al. Cerebrospinal Fluid Biomarkers and Clinical Progression in Patients with Subjective Cognitive Decline and Mild Cognitive Impairment. Journal of Alzheimer’s Disease 58, 939–950, doi:10.3233/JAD-161252 (2017).

